# Patient characteristics and predictors of mortality in 470 adults admitted to a district general hospital in England with Covid-19

**DOI:** 10.1101/2020.07.21.20153650

**Authors:** Joseph V Thompson, Nevan Meghani, Bethan M Powell, Ian Newell, Roanna Craven, Gemma Skilton, Lydia J Bagg, Irha Yaqoob, Michael J Dixon, Eleanor J Evans, Belina Kambele, Asif Rehman, Georges Ng Man Kwong

**Author notes:** **Corresponding author:** Joseph V Thompson BSc MChem MBBS MRCP, Department of Respiratory Medicine, Royal Oldham Hospital, Rochdale Road, Oldham, Greater Manchester, OL1 2JH, United Kingdom. **Alternate corresponding author:** Dr. Georges Ng Man Kwong, Department of Respiratory Medicine, Royal Oldham Hospital, Rochdale Road, Oldham, Greater Manchester, OL1 2JH, United Kingdom.

## Abstract

**Background:** Understanding risk factors for death in Covid-19 is key to providing good quality clinical care. Due to a paucity of robust evidence, we sought to assess the presenting characteristics of patients with Covid-19 and investigate factors associated with death.

**Methods:** Retrospective analysis of adults admitted with Covid-19 to Royal Oldham Hospital, UK. Logistic regression modelling was utilised to explore factors predicting death.

**Results:** 470 patients were admitted, of whom 169 (36%) died. The median age was 71 years (IQR 57–82), and 255 (54.3%) were men. The most common comorbidities were hypertension (n=218, 46.4%), diabetes (n=143, 30.4%) and chronic neurological disease (n=123, 26.1%). The most frequent complications were acute kidney injury (n=157, 33.4%) and myocardial injury (n=21, 4.5%). Forty-three (9.1%) patients required intubation and ventilation, and 39 (8.3%) received non-invasive ventilation Independent risk factors for death were increasing age (OR per 10 year increase above 40 years 1.87, 95% CI 1.57-2.27), hypertension (OR 1.72, 1.10-2.70), cancer (OR 2.20, 1.27-3.81), platelets <150×10^3^/µL (OR 1.93, 1.13-3.30), C-reactive protein ≥100 µg/mL (OR 1.68, 1.05-2.68), >50% chest radiograph infiltrates, (OR 2.09, 1.16-3.77) and acute kidney injury (OR 2.60, 1.64-4.13). There was no independent association between death and gender, ethnicity, deprivation level, fever, SpO_2_/FiO_2_ (oxygen saturation index), lymphopenia or other comorbidities.

**Conclusions:** We characterised the ‘first wave’ of patients with Covid-19 in one of England’s highest incidence areas, determining which factors predict death. These findings will inform clinical and shared decision making, including the use of respiratory support and therapeutic agents.

**Summary:** Increasing age, hypertension, cancer, platelets <150×10^3^/µL, CRP≥100 µg/mL, >50% chest radiograph infiltrates, and acute kidney injury predict in-hospital death from Covid-19, whilst gender, ethnicity, deprivation level, fever, SpO2/FiO2 (oxygen saturation index), lymphopenia and other comorbidities do not.

## Introduction

Coronavirus disease 2019 (Covid-19) has spread rapidly around the world from an initial cluster of viral pneumonia linked to a wet market in Wuhan, China in December 2019. The United Kingdom (UK) recorded its first case on 31^st^ January 2020 and as of 8^th^ July 2020 has recorded 44,321 deaths representing the 4^th^ highest per capita fatality rate worldwide at 66.6 per 100,000 people. [1]. Oldham, a large town forming part of the Greater Manchester urban conurbation, has recorded the fifth highest incidence rate nationally, with 783.0 cases per 100,000 people. [2]

Much of the data outlining patient characteristics and risk factors for severe disease and mortality has come from Chinese case series with unadjusted analyses, and it is unclear how these findings relate to other populations. Increasing age or SOFA score and D-dimer >1μg/mL were identified through multivariable analysis of 171 patients in Wuhan as independent predictors of death in hospital. [3] Other factors reported from China associated with severe disease or death include hypertension, cardiovascular disease, chronic lung disease [4], and chronic kidney disease [5]. These factors were also identified by Docherty et al as independent risk factors for death in an interim observational registry study of 20,133 hospitalised UK patients in addition to other chronic conditions. [6] Concern has been raised in the UK around the risk of death from Covid-19 in those of Black and Asian ethnicity, with these groups more likely to test positive and be hospitalised in the UK. [7]

Understanding factors that predispose to death from Covid-19 in hospital is crucial to providing good patient care; including clinical and shared decision making, timely escalation of care for respiratory support, and targeted use of potential antiviral or immunomodulatory therapies. As such, we sought to describe the characteristics and outcomes of those admitted with Covid-19 in Oldham, Greater Manchester, UK, including sociodemographic factors, presenting clinical, biochemical and radiological features, and complications. We assessed whether these characteristics were associated with death in hospital or within 30 days of discharge.

## Methods

### Study setting

The Royal Oldham Hospital is a 445 bed district general hospital located in Greater Manchester, a large urban conurbation in North West England serving a patient population of 230,000 people. [8] Oldham is amongst the most deprived areas in England, ranking 19^th^ of 317 local authorities nationally in the 2009 indices of multiple deprivation (IMD). [9] The predominant ethnic group is White (77.5%), with a considerably younger population of Pakistani (10.1%) and Bangladeshi (7.3%) heritage. [10] Population density is almost four times the national average at 15.8 persons per hectare. [11] Data was collected from electronic patient records in Symphony (EMIS Health) and HealthViews (Computer Sciences Corporation).

### Inclusion criteria

470 patients were included. Individuals ≥18 years admitted to Royal Oldham Hospital were included if SARS-2 CoV was detected by realtime polymerase chain reaction (PCR) from a nasopharyngeal swab prior to or during admission. Patients included were admitted between 12^th^ March 2020 and 19^th^ May 2020, in addition to those who acquired Covid-19 as inpatients.

### Outcome measures

The main outcome measure was death during admission or within 30 days of discharge, as recorded in the electronic patient record.

### Explanatory variables

Age, gender, ethnicity, postcode, date of admission, and date of discharge were recorded from structured fields. Age was categorised as <40 years, 40 – 50 years, 50 – 60 years, 60 – 70 years, 70 – 80 years, or > 90 years. Ethnicity was categorised into five groups – White British, Asian, African/Caribbean or other black background, Other white background, and other/not specified. Postcodes were used to generate IMD quintiles and deciles using the UK government Ministry of Housing, Communities and Local Government Postcode Lookup tool.

Physiological parameters at admission including heart rate, systolic blood pressure, respiratory rate, oxygen saturation, and temperature were collected from observations taken at triage in the emergency department. Fraction of inspired oxygen (FiO2) was estimated from oxygen flow rate using a standardised conversion table. SpO_2_ (oxygen saturation) / FiO_2_ ratio, a validated surrogate for PaO_2_ / FiO_2,_ was categorised as >315, <315, or <235 as indicators of mild, moderate, and severe acute lung injury (ALI) respectively. [12] Non-respiratory symptoms were categorised as gastrointestinal, cardiac, neurological, and/or other incidental symptoms. Clinical frailty scale (CFS) [13] was recorded from admission documentation. Immunosuppression was defined as recent chemotherapy or use of immunomodulatory medication including oral steroids on admission.

The following laboratory parameters were collected; admission lymphocyte count (x 10^3^/µL), lymphocyte nadir during admission (x 10^3^/µL), admission platelet count (x 10^3^/µL) and admission C-reactive protein (CRP) (µg/mL). Admission chest radiographs were categorised by degree of total new lung field infiltrates as none, less than 50%, or greater than 50%. Two clinicians reviewed radiographs where there was uncertainty.

Complications relating to Covid-19 were recorded, with pulmonary embolus, deep vein thrombosis and stroke recorded if confirmed by imaging studies. Super-added bacterial pneumonia was recorded if a causative organism was isolated. Acute kidney injury (AKI) was assessed using the Kidney Disease Improving Global Outcomes (KDIGO) criteria, and myocardial injury was deemed present if troponin elevation above the normal range (<2ng/L) was present without alternative explanation.

### Statistical analysis

Data was collected using Microsoft Excel. STATA 13 was used to produce summary statistics and assess associations with death in hospital or within 30 days of discharge.

For univariate analysis Pearson’s chi-squared test (*χ*^2^) was used to test the significance of categorical variables. Odds ratios for death and 95% confidence intervals were calculated for categorical variables. A K-sample equality-of-medians test was used to assess for significant differences in the medians of continuous variables for survivors compared to non-survivors given. A significance level of 0.05 was used for all statistical tests.

A logistic regression model to examine which factors predict death was constructed using dependent variables previously proposed as poor prognostic markers, as well as those with p values from univariate statistical tests of <0.25. The initial model was refined through backward elimination using the likelihood ratio test to assess for significant difference in model fit. CFS was removed from the model due to collinearity with age. The linktest program within STATA did not provide evidence of a model specification error. Goodness of fit for the final model deemed satisfactory using McFadden’s pseudo R squared test.

### Ethical approval

This study was approved by the Northern Care Alliance Research Management and Support team and did not require the approval from the NHS Health Research Authority.

## Results

### Sociodemographic factors

**Figure 1.**
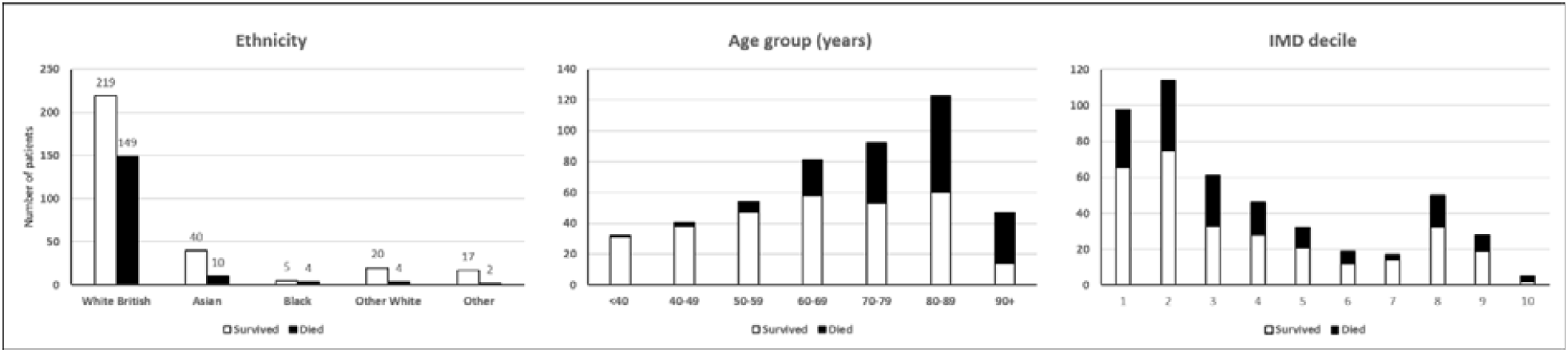
Sociodemographic characteristics of patients admitted with Covid-19 to Royal Oldham Hospital, UK stratified by survival to 30 days post-discharge. IMD – index of multiple deprivation, where the 1^st^ decile represents highest level of deprivation.

**Figure 2.**
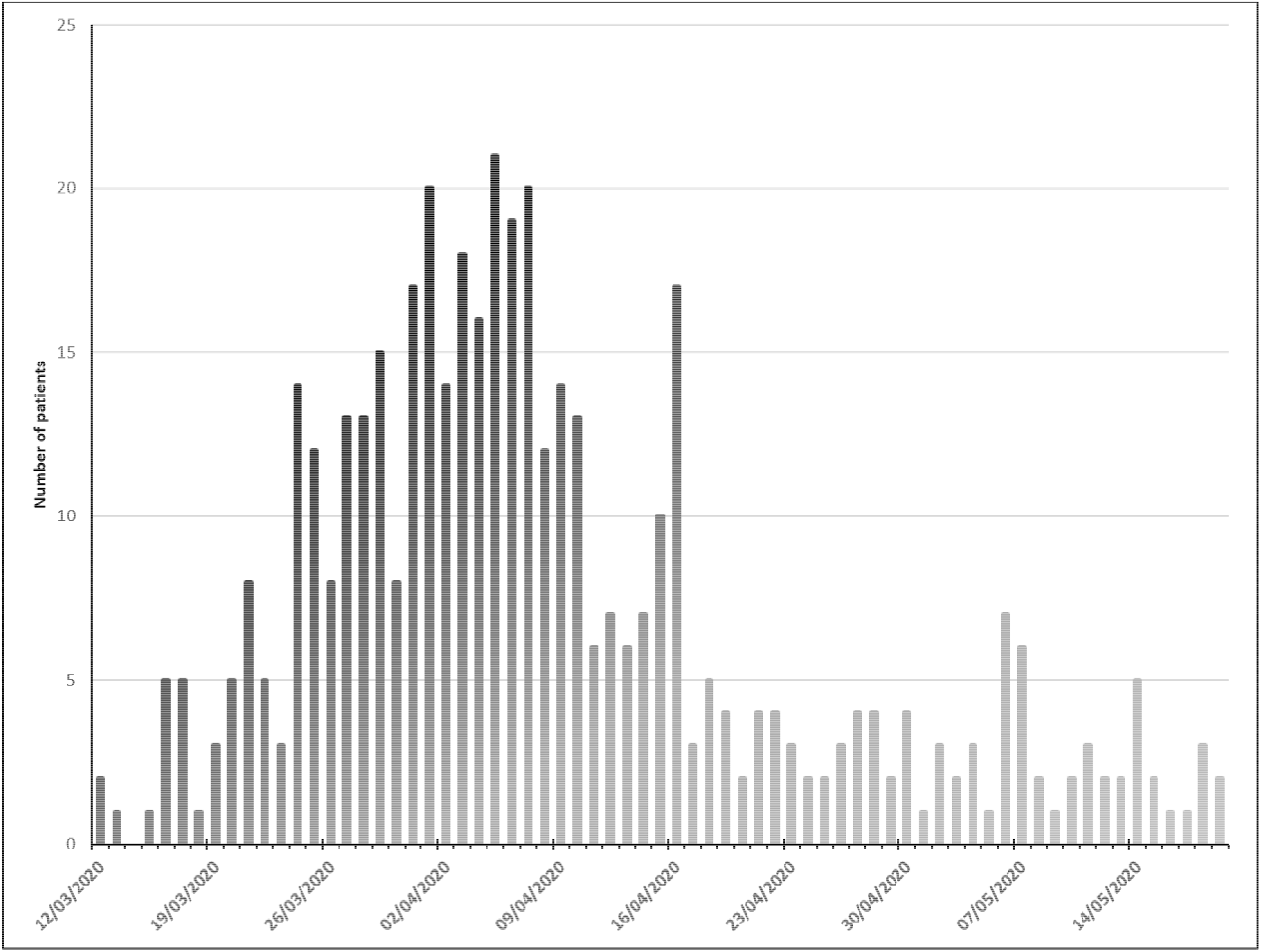
Patients admitted with Covid-19 to Royal Oldham Hospital, UK by date.

Of 470 patients admitted 36.0% (n=169) died during admission, within 30 days of discharge, or by the date of data collection. One patient remained an inpatient by the conclusion of data collection and 27 patients had been discharged from hospital <30 days. The median age of those admitted was 71 years (IQR 57–82 years), and the median age of non-survivors was 80 years. More men (54.3%, n=255) than women (45.7%, n=215) were admitted but following admission no difference in frequency of death was present (p=0.95). White British patients were the largest ethnic group (78.3%, n=368), followed by Asian (10.6%, n=50) and Other white background (5.1%, n=24), predominantly from Eastern Europe. In keeping with high local levels of deprivation, 44.9% of patients were classified as being in the most deprived quintile nationally. Death was significantly more frequent with increasing age (per 10 year age bracket increase above age 40 years OR 1.89, CI 1.62–2.20 p <0.001), and less frequent among those of Asian (OR 0.37, CI 0.18–0.76 p <0.01) and Other white background ethnicity (OR 0.29, CI 0.10–0.88 p <0.02) compared to White British patients. IMD quintile was not associated with death.

**Table 1.** Sociodemographic characteristics of patients admitted with Covid-19 stratified by survival

| **Sociodemographic characteristic** | **Total number of subjects (% of all patients)** | **Survival to 30 days post discharge** | **Death during admission or within 30 days of discharge** |
| --- | --- | --- | --- |
| **Total** | 470 (100%) | 301 (64.0%) | 169 (36.0%) |
| **Mean age (SD)** | 68.7 (17.4) | 63.4 (17.6) | 78.3 (12.1) |
| **Median age (IQR)** | 72 (57 - 82) | 65 (51 – 78) | 80 (72 – 87) |
| **Age group** |  |  |  |
| < 40 years | 32 (6.8%) | 31 (96.9%) | 1 (3.1%) |
| 40 – 49 years | 41 (8.7%) | 38 (92.7%) | 3 (7.3%) |
| 50 – 59 years | 54 (11.5%) | 47 (87.0%) | 7 (13.0%) |
| 60 – 69 years | 81 (17.2%) | 58 (71.6%) | 23 (28.4%) |
| 70 – 79 years | 92 (19.6%) | 53 (57.6%) | 39 (42.4%) |
| 80 – 89 years | 123 (26.1%) | 60 (48.8%) | 63 (51.2%) |
| ≥ 90 years | 47 (10.0%) | 14 (29.8%) | 33 (70.2%) |
| **Gender** |  |  |  |
| Female | 215 (45.7%) | 138 (64.2%) | 77 (35.8%) |
| Male | 255 (54.3%) | 163 (63.9%) | 92 (36.1%) |
| **Ethnicity** |  |  |  |
| White British | 368 (78.3%) | 219 (59.5%) | 149 (40.5%) |
| Asian | 50 (10.6%) | 40 (80.0%) | 10 (20.0%) |
| Black African, Caribbean or other black background | 9 (1.9%) | 5 (55.6%) | 4 (44.4%) |
| Other White Background | 24 (5.1%) | 20 (83.3%) | 4 (16.7%) |
| Other / Not specified | 19 (4.0%) | 17 (89.5%) | 2 (10.5%) |
| **IMD quintile** |  |  |  |
| 5 (Least deprived) | 33 (7.0%) | 21 (63.6%) | 12 (36.4%) |
| 4 | 67 (14.2%) | 46 (68.7%) | 21 (31.3%) |
| 3 | 51 (10.9%) | 33 (64.7%) | 18 (35.3%) |
| 2 | 108 (23.0%) | 62 (57.4%) | 46 (42.6%) |
| 1 (Most deprived) | 211 (44.9%) | 139 (65.9%) | 72 (34.1%) |

### Comorbidities

**Figure 3.**
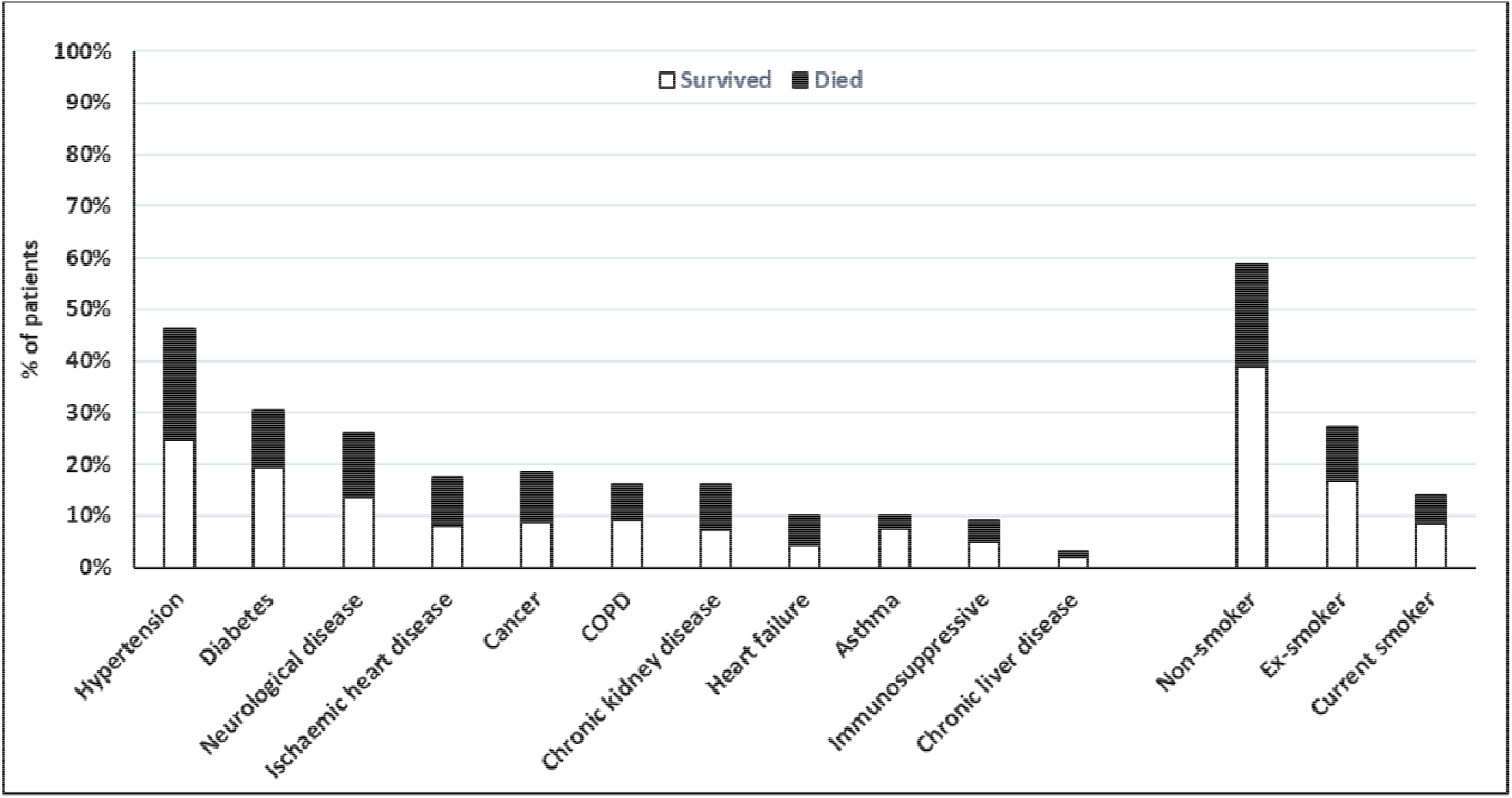
Comorbidities of patients admitted with Covid-19 to Royal Oldham Hospital, UK stratified by survival to 30 days post-discharge. COPD – chronic obstructive pulmonary disease.

Median CFS was significantly higher in non-survivors (6 IQR 4-7 vs. 3 IQR 2-5 for survivors), with a mean total number of significant comorbidities of 3.2 (SD 1.7) among those who died vs. 2.1 (SD 1.6) for survivors. 13.9% (n=66) had no prior significant medical conditions. The most common comorbidities were hypertension (46.4%, n = 218), diabetes (30.4%, n=143), and chronic neurological disease including dementia and other neurodegenerative conditions (26.1%, n=123). Hypertension, ischaemic heart disease, heart failure, chronic kidney disease, chronic neurological disease, and current or prior cancer were significantly more prevalent among those who died. Smoking, diabetes, chronic obstructive pulmonary disease, asthma, chronic liver disease, and immunosuppressive medication on admission were not significantly more common among non-survivors.

**Table 2.** Comorbidities of patients admitted with Covid-19 stratified by survival

| **Comorbidities** | **Total number of subjects (% of all patients)** | **Survival to 30 days post discharge** | **Death during admission or within 30 days of discharge** |
| --- | --- | --- | --- |
| **Median clinical frailty scale** | 4 (IQR 3-6) | 3 (IQR 2-5) | 6 (IQR 4-7) |
| **Mean total number of significant comorbidities** | 2.5 (SD 1.7) | 2.1 (SD 1.6) | 3.2 (SD 1.7) |
| **Smoking status** |  |  |  |
| **Non-smoker** | 276 (58.7%) | 183 (66.3%) | 93 (33.7%) |
| **Ex-smoker** | 128 (27.2%) | 79 (61.7%) | 49 (38.3%) |
| **Current smoker** | 66 (14.0%) | 39 (59.1%) | 27 (40.9%) |
| **Comorbidity** |  |  |  |
| **Hypertension** | 218 (46.4%) | 117 (53.7%) | 101 (46.3%) |
| **Diabetes** | 143 (30.4%) | 91 (63.6%) | 52 (36.4%) |
| **Ischaemic heart disease** | 81 (17.2%) | 38 (46.9%) | 43 (53.1%) |
| **Heart failure** | 47 (10%) | 20 (42.6%) | 27 (57.4%) |
| **COPD** | 76 (16.1%) | 43 (56.6%) | 33 (44.4%) |
| **Asthma** | 47 (10.0%) | 36 (76.6%) | 11 (23.4%) |
| **Chronic kidney disease** | 76 (16.1%) | 34 (44.7%) | 42 (55.3%) |
| **Chronic neurological disease** | 123 (26.1%) | 64 (52.0%) | 59 (48.0%) |
| **Chronic liver disease** | 14 (3.0%) | 9 (64.3%) | 5 (35.7%) |
| **Current or previous cancer** | 87 (18.5%) | 40 (46.0%) | 47 (54.0%) |
| **Current use of immunosuppressive medication** | 43 (9.1%) | 23 (53.5%) | 20 (46.5%) |

### Physiological parameters on admission

Tachypnoea was common, with 70% (n=329) presenting with a respiration rate ≥20/minute), and 38.5% (n=171) required supplemental oxygen. 15.7% (n=74) of patients were classified by SpO_2_/FiO_2_ index as severe ALI, 14.3% (n=67) as moderate ALI, and 70% (n=329) as mild or no ALI. Fever (temperature ≥37.8°C) was present in 40.6% (n=191) at triage. Admission respiration rate, severity of acute lung injury, heart rate, blood pressure, or fever were not associated with death.

The median duration of symptoms prior to presentation was 4 days (IQR 2-7), with no significant difference between survivors and non-survivors. Gastrointestinal symptoms (abdominal pain, nausea, vomiting or diarrhoea) were present in 18.9% (n=89) of patients, neurological symptoms (confusion or focal neurological signs) were present in 7.9% (n=37) patients, and presentations in keeping with cardiac disease (acute coronary syndrome, myocarditis) were evident in 1.3% (n=6) patients. Interestingly, one patient presented with posterior reversible encephalopathy syndrome, and one with cerebral vasculitis. 10.4% of patients (n=49) presented with clinical features consistent with alternative diagnoses in whom Covid-19 was found incidentally or diagnosed during admission.

**Table 3.** Clinical characteristics of patients admitted with Covid-19 stratified by survival

| **Clinical characteristic** | **Total number of subjects (% of all patients)** | **Survival to 30 days post discharge** | **Death during admission or within 30 days of discharge** |
| --- | --- | --- | --- |
| **Physiological parameters at triage** |  |  |  |
| **Median SpO2 / FiO2 index** | 442 (IQR 278–462) | 428 (IQR 475-457) | 443 (IQR 278-462) |
| **Median respiration rate** | 21 (IQR 19-26) | 20 (IQR 18-25) | 22(IQR 19-26) |
| **Median heart rate** | 90 (IQR 78-106) | 92 (IQR 79-107) | 90 (IQR 77-106) |
| **Median blood pressure** | 129 (IQR 115-146) | 131 (IQR 114-148) | 128 (IQR 116-146) |
| **No fever** | 279 (59.3%) | 177 (63.4%) | 102 (36.6%) |
| **Fever ($\geq 37.8^{\circ}\text{C}$)** | 191 (40.6%) | 124 (64.9%) | 67 (35.5%) |
| **Median duration of symptoms at presentation (days)** | 4 (IQR 2-7) | 5 (IQR 2-7) | 3 (IQR 1-7) |
| **GI symptoms (nausea, vomiting, diarrhoea, abdominal pain)** | 89 (18.9%) | 54 (60.7%) | 35 (39.3%) |
| **Neurological symptoms (confusion or focal neurological signs)** | 37 (7.9%) | 25 (67.6%) | 12 32.4%) |
| **Cardiac presentation (ACS/myocarditis)** | 6 (1.3%) | 4 (66.7%) | 2 (33.3%) |
| **Presentation consistent with possible Covid-19** | 421 (89.6%) | 265 (62.9%) | 156 (37.1%) |
| **Presentation not consistent with Covid-19<sup>a</sup>** | 49 (10.4%) | 36 (73.5%) | 13 (26.5%) |

<sup>a</sup>SARS-2 CoV identified incidentally at presentation or during admission.

### Biochemical and radiological features on admission

54.9% (n=258) of patients had a falling lymphocyte count during admission, with a median value on admission of 1.0 (IQR 0.7-1.4) and a median lymphocyte nadir of 0.7 (IQR 0.5-1.1). There was no significant difference in these values between survivors and non-survivors, and falling lymphocyte count during admission was not associated with death. The median admission CRP was significantly higher among those who died (94µg/mL IQR 38-167µg/mL vs. 75µg/mL IQR 34–132 µg/mL for survivors), whilst the median admission platelet count was significantly lower among non-survivors (190 x 10^3^/µL, IQR 142-275 x 10^3^/µL vs. 224 x 10^3^/µL, IQR 178-302 x 10^3^/µL for survivors). New infiltrates were noted on admission chest radiographs of 63.4% (n=298) of patients, with more than 50% total lung field involvement evident in 25.5% (n=120). Univariate analysis did not reveal an increased risk of death among those with new chest radiograph infiltrates.

**Table 4.**
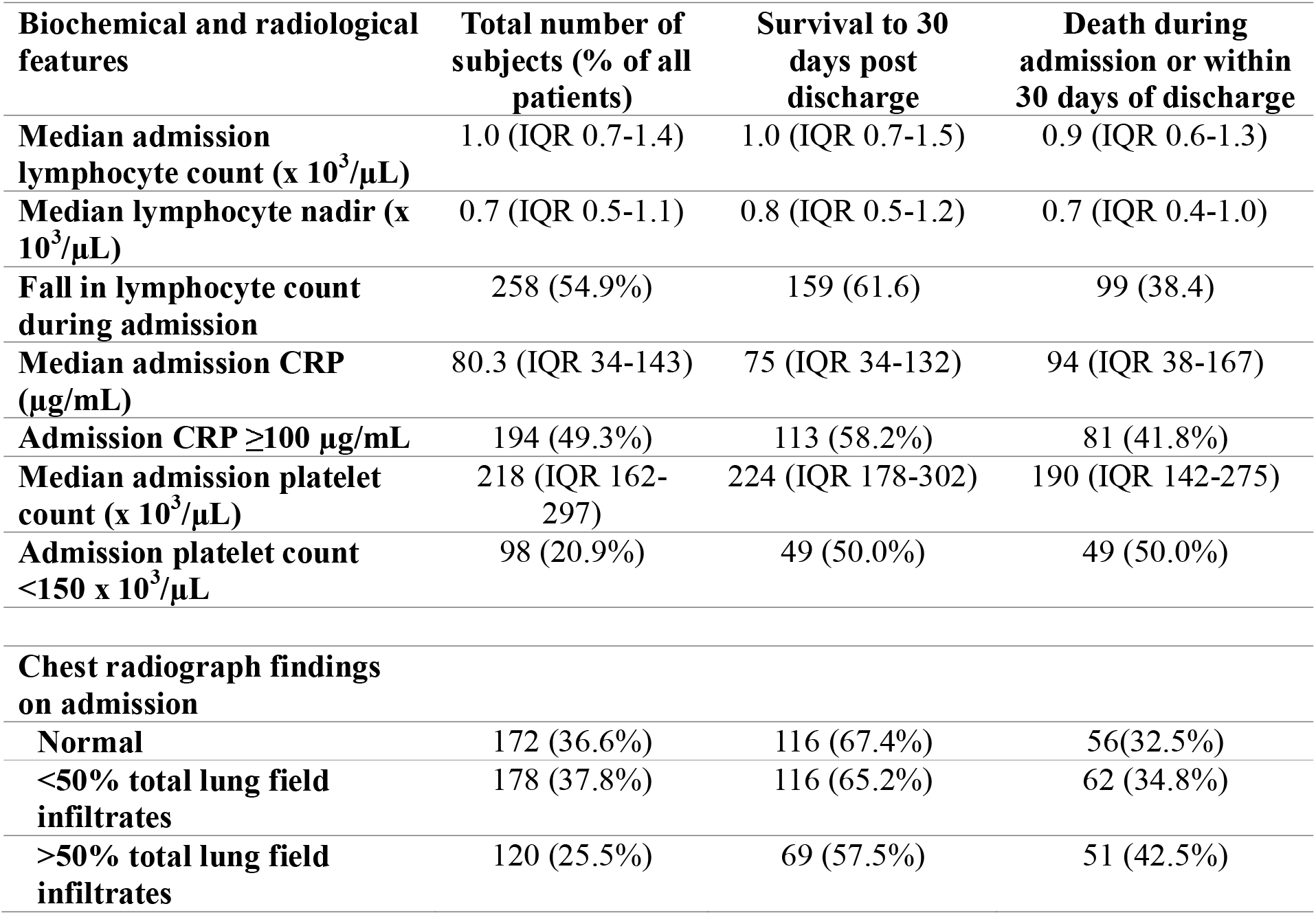
Biochemical and radiological characteristics of patients admitted with Covid-19 stratified by survival

| **Biochemical and radiological features** | **Total number of subjects (% of all patients)** | **Survival to 30 days post discharge** | **Death during admission or within 30 days of discharge** |
| --- | --- | --- | --- |
| **Median admission lymphocyte count (x 10<sup>3</sup>/μL)** | 1.0 (IQR 0.7-1.4) | 1.0 (IQR 0.7-1.5) | 0.9 (IQR 0.6-1.3) |
| **Median lymphocyte nadir (x 10<sup>3</sup>/μL)** | 0.7 (IQR 0.5-1.1) | 0.8 (IQR 0.5-1.2) | 0.7 (IQR 0.4-1.0) |
| **Fall in lymphocyte count during admission** | 258 (54.9%) | 159 (61.6) | 99 (38.4) |
| **Median admission CRP (μg/mL)** | 80.3 (IQR 34-143) | 75 (IQR 34-132) | 94 (IQR 38-167) |
| **Admission CRP ≥100 μg/mL** | 194 (49.3%) | 113 (58.2%) | 81 (41.8%) |
| **Median admission platelet count (x 10<sup>3</sup>/μL)** | 218 (IQR 162-297) | 224 (IQR 178-302) | 190 (IQR 142-275) |
| **Admission platelet count <150 x 10<sup>3</sup>/μL** | 98 (20.9%) | 49 (50.0%) | 49 (50.0%) |
| **Chest radiograph findings on admission** |  |  |  |
| **Normal** | 172 (36.6%) | 116 (67.4%) | 56(32.5%) |
| **<50% total lung field infiltrates** | 178 (37.8%) | 116 (65.2%) | 62 (34.8%) |
| **>50% total lung field infiltrates** | 120 (25.5%) | 69 (57.5%) | 51 (42.5%) |

### Complications and respiratory support

The most commonly observed complications due to Covid-19 were acute kidney injury (AKI) (33.4% n=157), myocardial injury (4.5% n=21), and bacterial pneumonia (3.2%, n=15). Pulmonary embolus was identified on 5 of 24 computed tomography pulmonary angiograms (1.1% of all patients), whilst deep vein thrombosis was identified from 8 of 19 duplex venous ultrasounds (1.7% of all patients). Acute kidney injury and myocardial injury were more common among those who died.

Respiratory support was required by 14.7% (n=69) of patients, with 1.7% (n=8) of patients receiving High flow nasal cannula (HFNC), 8.3% (n=39) receiving non-invasive continuous positive pressure airway ventilation (CPAP), and 9.2% (n=43) intubated and mechanically ventilated (IMV). The use of more than one mode of respiratory support was frequently employed.

**Table 5.** Complications and respiratory support for patients admitted with Covid-19 stratified by survival.

| **Respiratory support and complications** | **Total number of subjects (% of all patients)** | **Survival to 30 days post discharge** | **Death during admission or within 30 days of discharge** |
| --- | --- | --- | --- |
| **Mode of respiratory support** |  |  |  |
| **CPAP** | 39 (8.3%) | 26 (66.7%) | 13 (33.3%) |
| **HFNC** | 8 (1.7%) | 3 (37.5%) | 5 (62.5%) |
| **IMV** | 43 (9.1%) | 28 (65.1%) | 15 (34.9%) |
| **Complications during admission** |  |  |  |
| **Pulmonary embolism** | 5 (1.1%<sup>a</sup>) | 4 (80.0%) | 1 (20.0%) |
| **Deep vein thrombosis** | 8 (1.7%<sup>b</sup>) | 5 (62.5%) | 3 (37.5%) |
| **Myocardial Injury** | 21 (4.5%) | 9 (42.9%) | 12 (57.1%) |
| **Acute kidney injury** | 157 (33.4%) | 72 (45.8%) | 85 (54.1%) |
| **Stroke** | 1 (0.2%) | 1 (100%) | 0 (0%) |
| **Bacterial pneumonia** | 15 (3.2%) | 12 (80.0%) | 3 (20.0%) |

<sup>a</sup> 1.1% of all patients, 20.8% of those who underwent CTPA
<sup>b</sup> 1.7% of all patients, 42.1 of those who under venous duplex ultrasound

### Multivariate analysis

Logistic regression identified the following variables which were also significant in univariate analysis as independent risk factors for death - increasing age, history of hypertension, current or previous cancer, admission CRP ≥100µg/mL, admission platelet count <150 x 10^3^ /µL, and AKI. Additionally, the model identified >50% total lung field infiltrates, which was not significant in univariate analysis, as a significant risk factor for death. Ethnicity, history of chronic kidney disease, ischaemic heart disease, heart failure or chronic neurological disease, and myocardial injury, which were observed more frequently among those who died, were not significant in the adjusted model. No other explanatory variables were found to be significant independent predictors of death.

**Table 6.** Significant risk factors for death among patients admitted with Covid-19 – univariate and multivariate logistic regression

| **Risk factor** | **Univariate odds ratio for death compared to reference group** | **95% confidence interval** | **p value (Wald test)** | **Adjusted odds ratio for death compared to reference group (Logistic regression)** | **95% confidence interval** | **p value (Wald test)** |
| --- | --- | --- | --- | --- | --- | --- |
| **Age - per 1 year increase** | 1.07 | 1.05–1.08 | <0.001 |  |  |  |
| **Age - per 10 year increase above age 40** | 1.89 | 1.62–2.20 | <0.001 | 1.87 | 1.57-2.27 | <0.001 |
| **Ethnicity** |  |  |  |  |  |  |
| **White British** | 1.00 |  |  |  |  |  |
| **Asian** | 0.37 | 0.18–0.76 | <0.01 |  |  |  |
| **Black African, Caribbean or other black background** | 1.18 | 0.31–4.45 | 0.81 |  |  |  |
| **Other White Background** | 0.29 | 0.10–0.88 | 0.03 |  |  |  |
| **Other / Not specified** | 0.17 | 0.04–0.76 | 0.02 |  |  |  |
| **Comorbidities** |  |  |  |  |  |  |
| **Mean clinical frailty scale \*** | 1.72 | 1.52–1.94 | <0.001 | Not included in regression model due to collinearity with age. |  |  |
| **Total number of significant comorbidities \*** | 1.45 | 1.31–1.67 | <0.001 |  |  |  |
| **Hypertension** | 2.34 | 1.49–3.43 | <0.001 | 1.72 | 1.10–2.70 | 0.02 |
| **Chronic kidney disease** | 2.60 | 1.56–4.28 | <0.001 |  |  |  |
| **Ischaemic heart disease** | 2.36 | 1.45–3.84 | <0.01 |  |  |  |
| **Heart failure** | 2.67 | 1.45–4.93 | <0.01 |  |  |  |
| **Chronic neurological disease** | 1.99 | 1.31–3.02 | <0.001 |  |  |  |
| **Cancer – current or** | 2.51 | 1.57–4.04 | <0.001 | 2.20 | 1.27–3.81 | <0.01 |
| **previous** |  |  |  |  |  |  |
| **Biochemical** |  |  |  |  |  |  |
| Admission CRP $\geq 100$ $\mu\text{g/mL}$ | 1.53 | 1.05–2.24 | 0.03 | 1.68 | 1.05–2.68 | 0.03 |
| Admission platelet count $<150 \times 10^3/\mu\text{L}$ | 2.10 | 1.34–3.30 | $<0.001$ | 1.93 | 1.13–3.30 | 0.01 |
| **Admission chest radiograph** |  |  |  |  |  |  |
| Normal | 1.00 |  |  |  |  |  |
| $<50\%$ total lung field infiltrates | 1.10 | 0.71–1.73 | 0.65 | | | |
| $>50\%$ total lung field infiltrates | 1.53 | 0.95–2.48 | 0.08 | 2.09 | 1.16–3.77 | 0.01 |
| **Complications** |  |  |  |  |  |  |
| Myocardial injury | 2.48 | 1.02–6.01 | 0.04 |  |  |  |
| AKI | 3.20 | 2.15–4.81 | $<0.001$ | 2.60 | 1.64–4.13 | $<0.001$ |

\*per 1 unit increase

**Figure 4.**
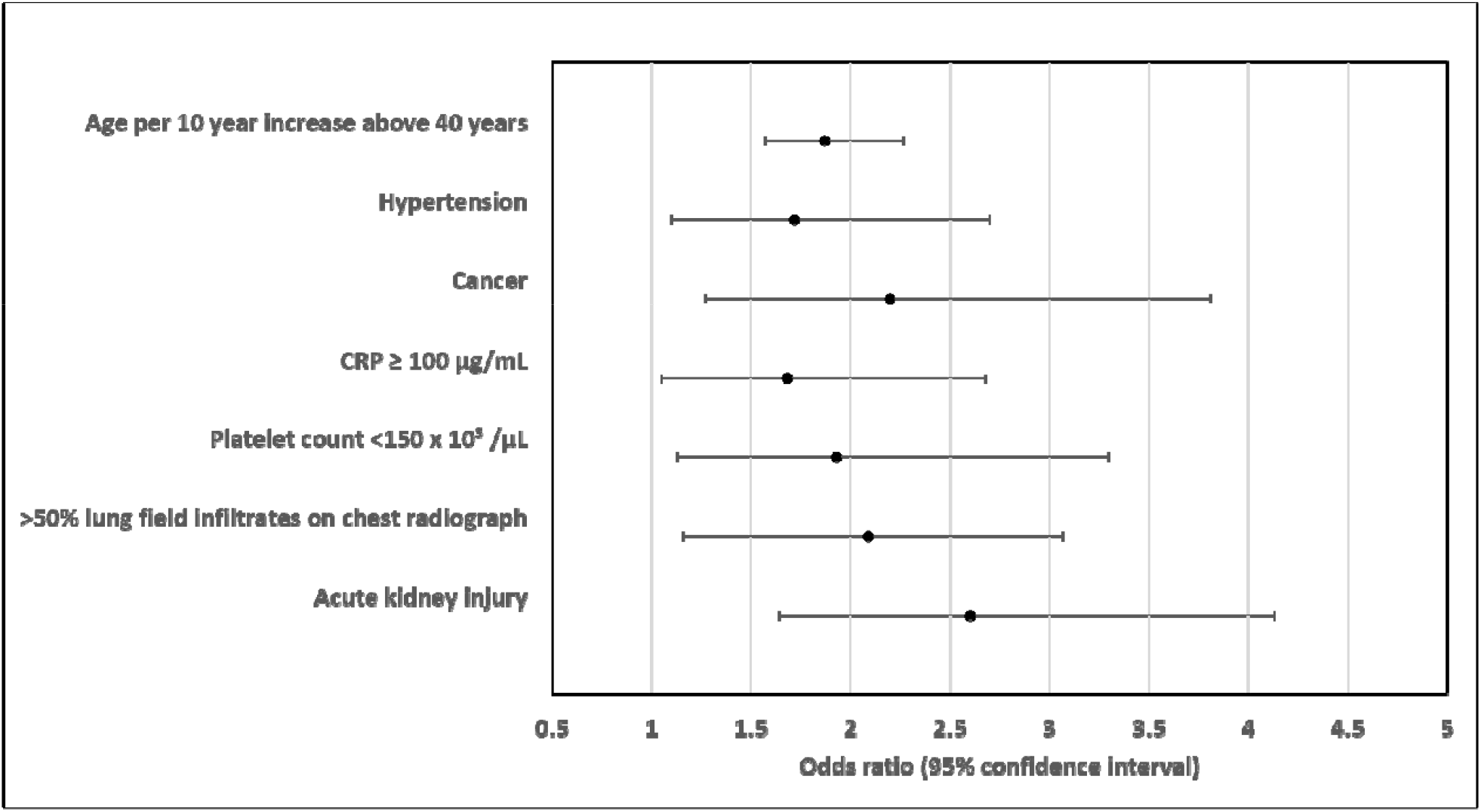
Independent predictors of death for patients admitted with Covid-19 to Royal Oldham Hospital, UK. All risk factors assessed on admission to hospital with the exception of acute kidney injury which was classified as a complication of Covid-19 and recorded if present at any time during admission.

## Discussion

### Summary of main findings and comparison to other studies

We assessed a wider range of potential risk factors for death than previous studies, with selection of variables guided by their clinical importance to us as hospital physicians in describing the overall clinical state of patients admitted to hospital with acute illness. In keeping with prior studies which adjusted for either no or fewer clinically relevant factors [3, 4, 6], we identified a wide range of factors that were more common among non-survivors. However, only increasing age, hypertension, previous or current cancer, platelet count <150 x 10^3^/µL, CRP ≥100 µg/mL, greater than 50% total lung field infiltrates on chest radiograph, and AKI were identified as independent risk factors for death in our clinically relevant and statistically robust regression model. Our mortality rate of 36.0% (n=169) is in keeping with the UK’s mortality rate for those hospitalised with Covid-19. [14]

More men than women were admitted, although the 53.2% of men in our cohort is significantly smaller than the 60% of males admitted in a previous large UK multicentre study. [6] Incredibly, we observed no difference in mortality by gender, despite men accounting for 80% of those requiring intubation, and gender remained a non-significant factor following multivariate analysis. This finding is in stark contrast to reports that men hospitalised with Covid-19 are at higher risk of death. [6] Whilst death was more frequent among White British patients, no ethnic group was an independent risk factor for mortality. Ethnicity was strongly attenuated in the logistic regression model by age, with the median age of White British patients 20 and 31 years greater than those of Black and Asian patients respectively. This study was underpowered to detect a mortality difference in Black patients given the low number admitted.

Hypertension, diabetes, cancer, and current smoking were recorded far more frequently in our cohort than in the largest UK study assessing the characteristics of those admitted with Covid-19. [6] Hypertension was present in almost half of those admitted and 46.3% (n=101) of this group died suggesting patients with hypertension should be regarded as a particularly high-risk group. Whilst hypertension and cancer were independent risk factors for death in our multivariate analysis, [6] previously reported factors including diabetes, chronic obstructive pulmonary disease (COPD), and ischaemic heart disease [3, 15] were not found to be independent risk factors. The majority of patients admitted had at least one comorbidity, and only four patients without comorbidity died, suggesting that those without significant medical problems are at very low risk of death.

Lymphopenia was observed in 63.2% of patients, in keeping with previous studies suggesting it is an important predictor of severe disease. [16] However, we did not find any evidence that admission lymphocyte count, lymphocyte nadir, or falling lymphocytes are associated with death in either univariate or multivariate analysis, suggesting that any prognostic implication is rendered prior to the point of hospital admission. We suggest that lymphocyte count is of diagnostic utility only and is not a useful prognostic marker during hospital admission. However, we identified CRP ≥100 µg/mL and platelet count <150 x 10^3^/µL as independent risk factors for death, which could be monitored serially to risk stratify patients.

SpO_2_/FiO_2_ is a validated surrogate for PaO_2_/FiO_2_ used to classify severity of acute respiratory distress syndrome (ARDS), [12] a condition thought to share pathophysiological mechanisms with Covid-19. Surprisingly, no difference in SpO_2_/FiO_2_ was observed between survivors and non-survivors, but the presence of infiltrates involving more than 50% of the lung fields on admission chest radiograph was an independent predictor of death. We recommend greater prognostic weight be given to severity of radiological changes than SpO_2_ and oxygen requirement at presentation.

Optimal fluid management is imperative given the high incidence of acute kidney injury and independent association with death in our cohort, underlining a recent recommendation to maintain euvolaemia from the UK National Institute for Health and Care Excellence (NICE) [17]. This followed initial concerns that fluid therapy may increase mortality through worsening of non-cardiogenic pulmonary oedema. It is unclear how changing practice has impacted upon the incidence of AKI and its attributable mortality.

### Strengths

We assessed variables available on admission that we as clinicians felt were important in describing the clinical state of acutely unwell patients. Our results are of particular relevance to areas of similar sociodemographic makeup including cities in the UK, and overseas. Unlike previous studies, we report near complete outcome data for our cohort, with one patient hospitalised at conclusion of data collection. We utilised an iterative process (backward elimination) to derive a fully adjusted model with good predictive ability.

### Limitations

We assessed risk factors for death present in hospitalised patients and have not investigated those not requiring admission. Consequently, there may be risk factors that confer risk for hospitalisation, and therefore a higher risk of death, that we have not identified. This is suggested, for example, by the disproportionate number of men admitted.

## Conclusions

We characterised the ‘first wave’ of patients presenting to a district general hospital with Covid-19 in one of England’s highest incidence areas, and determined that increasing age, hypertension, cancer, CRP >100µg/mL, platelets <150 x 10^3^/µL, severe chest radiograph appearance and acute kidney injury independently predict death following admission. We propose greater prognostic weight be given to these factors than to SpO2, oxygen requirement or lymphopenia, which did not predict death. Results from both our unadjusted and adjusted analyses dispute other previously proposed risk factors for death in hospital including male gender and diabetes.

These findings have the potential to inform decision making by front-line clinicians at the point of admission to hospital allowing risk stratification, improved shared decision-making with patients, the timely provision of respiratory support and targeted use of potential therapies for those most likely to benefit. If our model is prospectively validated, we anticipate introducing a clinical decision tool to aid prognostication for hospitalised patients with Covid-19.

## Data Availability

Data may be available from the authors upon request, dependent on institutional approval.

